# Comparative Efficacy of Brush Cytology and Forceps Biopsy in the Diagnosis of Malignant Biliary Strictures: A Single-Center Study in Pakistan

**DOI:** 10.1101/2024.08.03.24311458

**Authors:** Abdullah Khalid, Sara Qureshi, Imran Ali Syed, Hassan Nadeem, Ahmad Karim Malik, Usman Iqbal Aujla

**Affiliations:** Gastroenterology and Hepatology Department, Pakistan Kidney and Liver Institute & Research Center, DHA Phase VI, Lahore, Pakistan

**Keywords:** Biliary Strictures, Brush Cytology, Forceps Biopsy, Diagnostic Efficacy, Malignancy Detection

## Abstract

**Introduction:** The accurate diagnosis of biliary strictures or narrowing of the bile duct is challenging in clinical practice. Differentiating malignant from benign strictures is critical, because these treatments differ considerably. Although noninvasive imaging techniques help identify these strictures, they do not provide definitive tissue diagnosis, making techniques such as brush cytology and forceps biopsy essential. This study aimed to address the dearth of comparative research on the diagnostic efficacy of brush cytology and forceps biopsy in Pakistan.

**Methods:** This single-center observational study was conducted at the Pakistan Kidney and Liver Institute & Research Center (PKLI&RC), Lahore, from March 2019 to January 2023. Patients with clinically and radiologically suspected biliary strictures were included in the study. Both brush cytology and forceps biopsy samples were subjected to cytopathological analysis by blinded pathologists. Sensitivity, specificity, and predictive values were among the key metrics analyzed.

**Results:** The study included 54 patients in the biopsy group and 89 in the brushing group. In terms of diagnostic metrics, the biopsy technique displayed a sensitivity of 65.8%, specificity of 92.3%, positive predictive value (PPV) of 96.4%, and accuracy rate of 72.2%. For the brushing technique, the sensitivity, specificity, PPV, and accuracy were 56.7 %, 93.3%, 83.1%, and 62.9%, respectively. Although both methods showed high specificity, the biopsy technique exhibited a slightly better sensitivity and overall accuracy.

**Conclusion:** Our findings underscore the importance of the biopsy method in the accurate diagnosis of malignant biliary strictures, showing marginally superior sensitivity and overall accuracy compared to brush cytology. This knowledge can guide clinicians in Pakistan and similar settings in making informed diagnostic choices to improve patient outcomes.

## INTRODUCTION

Biliary strictures represent fixed narrowing within a segment of the bile duct, leading to proximal biliary dilatation and obstructive jaundice symptoms [1]. The accurate diagnosis of these strictures remains a challenge in clinical practice. They often arise from underlying conditions, including malignancies in the proximal and distal bile ducts, hepatic ducts, pancreas, gallbladder, or near the ampulla of Vater [2].

Differentiating between malignant and benign strictures based on imaging is a daunting task. Benign conditions may imitate a malignant stricture appearance, making sole reliance on clinical evaluation and imaging a potential source of misdiagnosis [3]. While non-invasive techniques, such as CT, Ultrasound, and MRI Cholangiography, are beneficial in identifying these strictures, they fail to provide a definitive tissue diagnosis [4]. Tissue confirmation is pivotal, especially when a malignant stricture is suspected.

A proper diagnosis of malignant biliary strictures cannot be overemphasized. The treatment varies significantly between malignant and benign strictures. Malignancies typically require aggressive measures, from surgery to radiation, each with morbidity and mortality rates [5]. Conversely, benign strictures can often be managed using minimally invasive procedures, such as endoscopy [6]. Therefore, an accurate diagnosis is paramount to ensure that patients receive optimal tailored treatment without unnecessary delays or interventions.

The current diagnostic arsenal for malignant strictures includes brush cytology, forceps biopsy, intraductal bile aspiration cytology, and cytopathological analysis [5,7]. While brush cytology stands out because of its accessibility and safety profile, forceps biopsy presents challenges, particularly when biliary sphincterotomy is required [8]. Despite its complexity, forceps biopsy has a significant diagnostic advantage, especially for focal strictures [9]. However, risks, such as pancreatitis, bleeding, and perforation, are associated with this method.

Clinicians and patients share the universal objective of improving the management of biliary strictures. Given the inherent challenges in distinguishing between malignant and benign strictures, an accurate pathological diagnosis remains at the forefront of this objective.

Despite its clinical significance, Pakistan lacks in-depth research comparing brush cytology and forceps biopsy efficiency. This study bridges this knowledge gap by targeting the local population. By analyzing metrics such as sensitivity, specificity, and predictive values of both techniques, we aimed to offer insights into better treatment strategies, ultimately elevating patient care.

## METHODS

This single-center observational study was conducted at the Pakistan Kidney and Liver Institute & Research Center (PKLI&RC), located in Lahore, Pakistan, between March 2019 and January 2023. Ethical approval was obtained from the Institutional Review Board of PKLI&RC. The study population comprised patients who were clinically and radiologically suspected to have biliary strictures.

Patient inclusion was based on the following criteria: age > 18 years, radiological evidence suggestive of biliary strictures, and consent for both brush cytology and forceps biopsy for diagnostic evaluation. Patients with a known history of coagulopathy, those who underwent sphincterotomy, or those who declined to participate in the study were excluded. All procedures were performed using endoscopic retrograde cholangiopancreatography (ERCP). For brush cytology, a specialized cytology brush was passed through the endoscope into the bile duct to ensure that cells from the suspicious lesion were adequately sampled. Biopsy forceps were advanced through the endoscope and used to extract a tissue sample from the stricture site.

The samples obtained were sent to the pathology department of the hospital. Both brush cytology and forceps biopsy samples were subjected to cytopathological analysis by experienced pathologists who were blinded to the clinical details of the patients. The diagnostic metrics analyzed were the sensitivity, specificity, positive predictive value, negative predictive value, and overall accuracy. The gold standard for diagnosis is a combination of clinical, radiological, and histopathological findings and, in some cases, surgical outcomes.

Statistical analyses were performed using SPSS version 25. Descriptive statistics were computed for all variables. The diagnostic efficacies of brush cytology and forceps biopsy were compared using the chi-square test for categorical variables, with a p-value of less than 0.05, which was considered statistically significant. Furthermore, confidence intervals (95% CI) were determined for the sensitivity, specificity, and predictive values.

## RESULTS

In our retrospective single-center analysis conducted at the Pakistan Kidney and Liver Institute & Research Center (PKLI&RC) in Lahore, we performed a meticulous comparison between the diagnostic efficacies of brush cytology and forceps biopsy for detecting malignant biliary strictures. Spanning the period from March 2019 to January 2023, this research delved into the specific strengths and limitations of both diagnostic modalities, targeting a local Pakistani population. Drawing upon a cohort of patients exhibiting clinical and radiological signs of biliary strictures, this study aimed to identify the most reliable diagnostic tool by evaluating key metrics, such as sensitivity, specificity, and predictive values.

### Patient Demographics and Preoperative Characteristics

The study cohort comprised 54 patients in the biopsy group and 89 in the brushing group. The mean age of the biopsy cohort was 52.8 ± 14.8 years, and that of the brushing cohort was 55.7 ± 13.7 years. The sex distribution was almost equal in the biopsy cohort (male/female:27/27), whereas in the brushing cohort, there were more males (60/29). In terms of stricture sites, the majority of patients from both cohorts underwent CBD biopsies (59.2% for biopsy and 62.9% for brushing). Malignant strictures were identified in 33.3% and 48.3% of the patients in the biopsy and brushing cohorts, respectively. The mean CA 19-9 (U/mL) level was 492.3 ± 1726.8 in the biopsy group and 757.9 ± 1930.7 in the brushing group (**Table 1**).

**Table 1.** Patient Demographics and Preoperative Characteristics.

| Characteristic | Biopsy Cohort (n=54) | Brushing Cohort (n=89) |
| --- | --- | --- |
| Age | 52.8 ± 14.8 | 55.7 ± 13.7 |
| Gender (male/female) | 27/27 | 60/29 |
| CA 19-9 (U/mL) | 492.3 ± 1726.8 | 757.9 ± 1930.7 |
| **Site** |  |  |
| CBD Biopsy | 32 (59.2%) | 56 (62.9%) |
| Biliary Biopsy | 18 (33.3%) | 31 (34.8%) |
| CHD Biopsy | 4 (7.4%) | 1 (1.1%) |
| **Strictures** |  |  |
| Benign | 36 (66.6%) | 46 (51.7%) |
| Malignant | 18 (33.3%) | 43 (48.3%) |

CA 19-9 = Carbohydrate Antigen 19-9; U/mL = Units per mL; CBD, common bile duct; CHD, common hepatic duct.

### Distribution of Diagnoses Pre and Post-ERCP

In the biopsy cohort, before ERCP, there were six cases of benign biliary strictures, which increased to 10 post-ERCP. The number of patients diagnosed with cholangiocarcinoma was 25, which reduced to 13 post-ERCP. In the brushing cohort, the number of benign biliary stricture cases increased from 7 to 13 post-ERCP. Similarly, the number of cholangiocarcinoma cases decreased from 42 to 21 post-ERCP. Several other diagnostic changes were observed after ERCP in both benign and malignant categories (**Tables 2** and **3**).

**Table 2.**
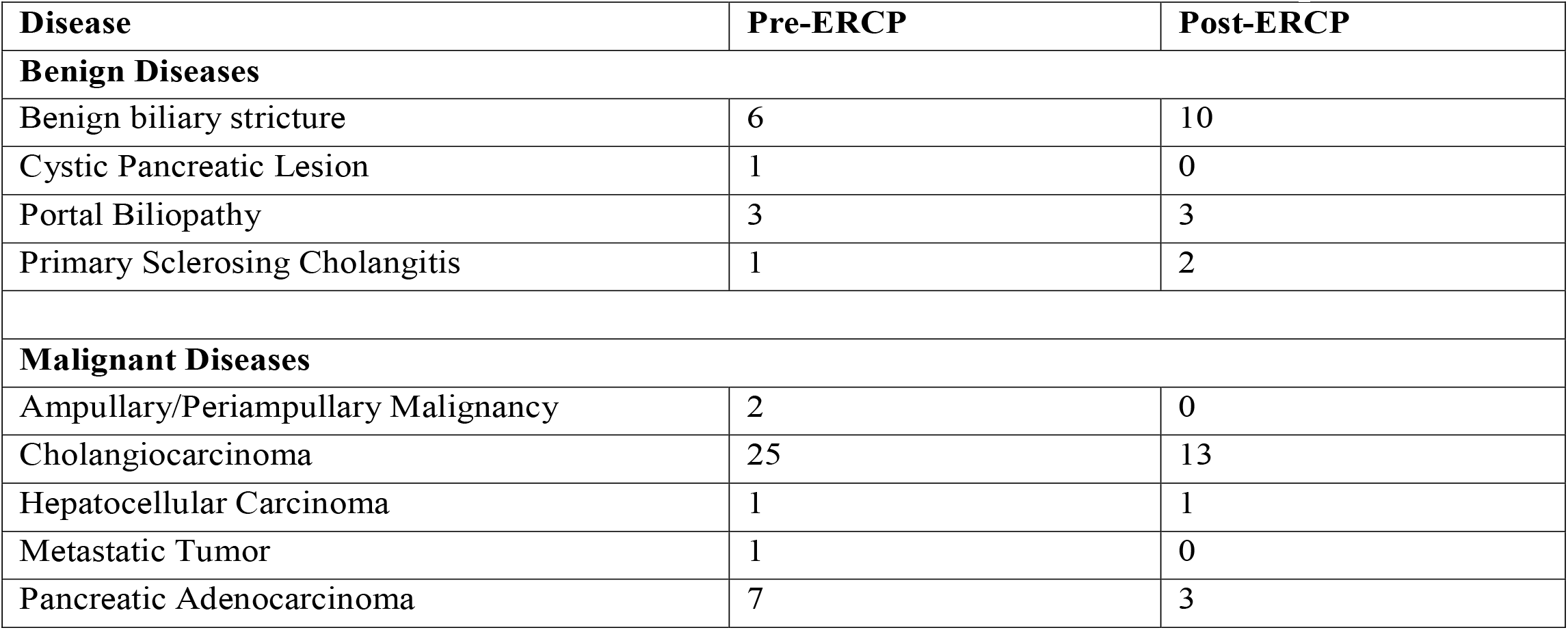
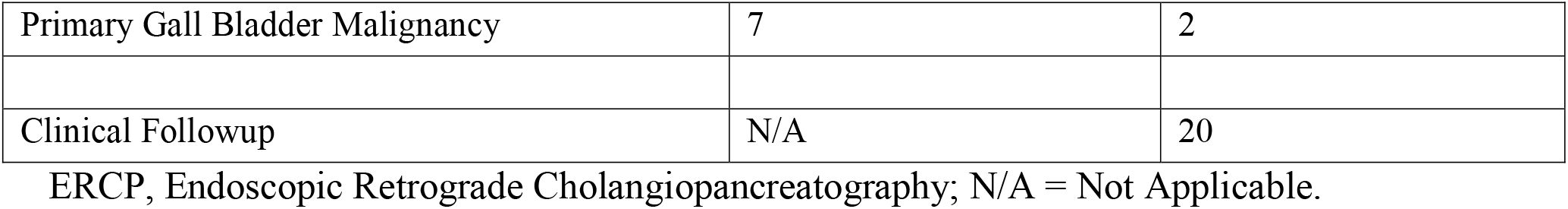
Distribution of Diagnoses Pre- and Post-ERCP for Biopsy Cohort (n=54).

| Disease | Pre-ERCP | Post-ERCP |
| --- | --- | --- |
| **Benign Diseases** |  |  |
| Benign biliary stricture | 6 | 10 |
| Cystic Pancreatic Lesion | 1 | 0 |
| Portal Biliopathy | 3 | 3 |
| Primary Sclerosing Cholangitis | 1 | 2 |
| **Malignant Diseases** |  |  |
| Ampullary/Periampullary Malignancy | 2 | 0 |
| Cholangiocarcinoma | 25 | 13 |
| Hepatocellular Carcinoma | 1 | 1 |
| Metastatic Tumor | 1 | 0 |
| Pancreatic Adenocarcinoma | 7 | 3 |
| Primary Gall Bladder Malignancy | 7 | 2 |
| Clinical Followup | N/A | 20 |

ERCP, Endoscopic Retrograde Cholangiopancreatography; N/A = Not Applicable.

**Table 3.**
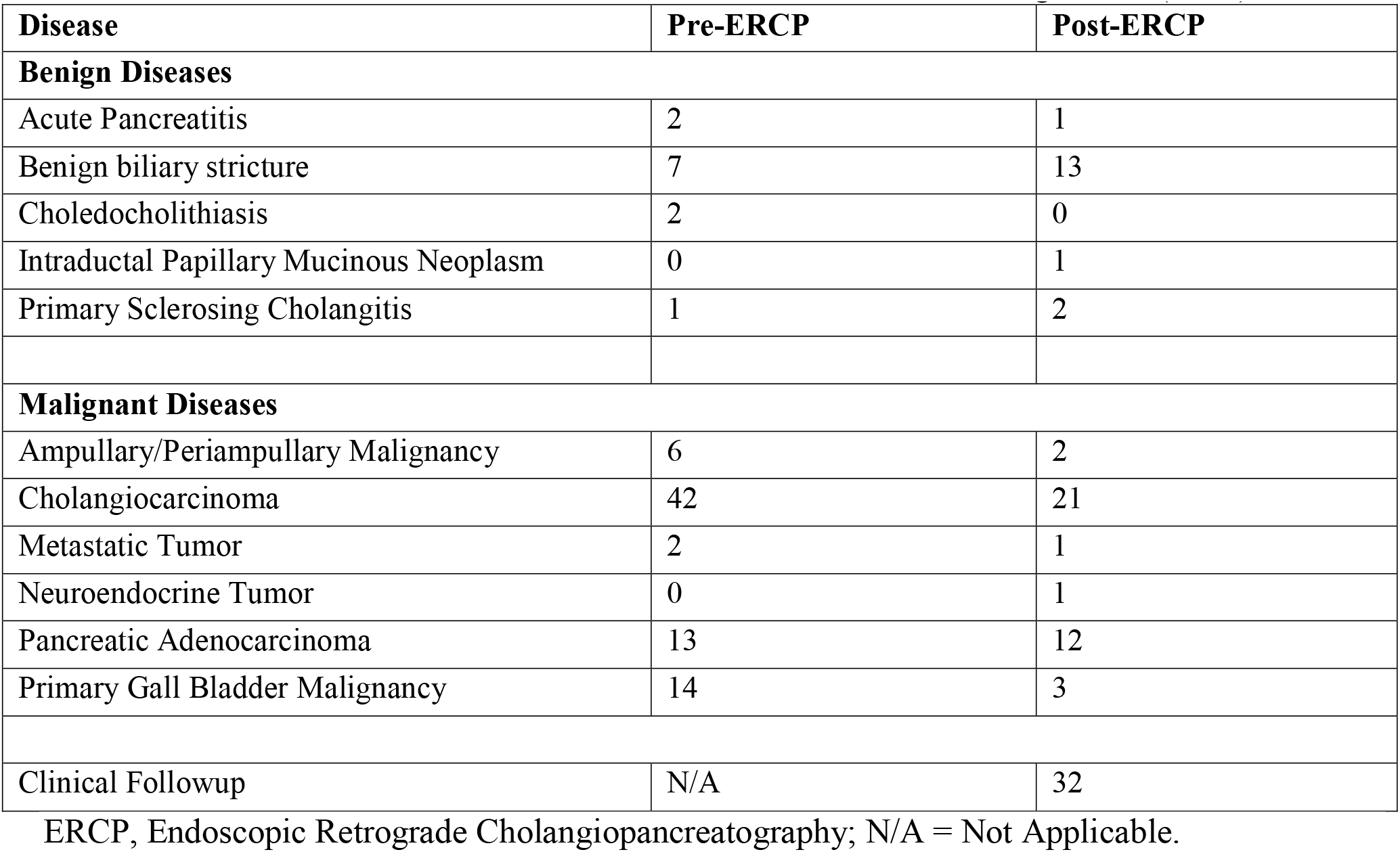
Distribution of diagnoses before and after ERCP in the brushing cohort (n=89).

### Diagnostic Efficacy of Techniques

In terms of diagnostic metrics, the biopsy technique demonstrated a sensitivity of 65.8% (95% CI:49.4-79.9%), specificity of 92.3% (95% CI:63.9-99.8%), positive predictive value (PPV) of 96.4% (95% CI:81.6-99.9%), and an accuracy rate of 72.2% (95% CI:58.3-83.5%). The negative predictive value (NPV) for biopsy was 46.1% (95% CI:26.5-66.6%).

For the brushing technique, the sensitivity was 56.7% (95% CI:44.7-68.2%), specificity was 93.3% (95% CI:68.0-99.8%), PPV was 83.1% (95% CI:73.7-90.2%), and accuracy was 62.9% (95% CI:52.0-72.9%). The NPV for brushing was 30.43% (95% CI:17.7-45.7%) (**Table 4**).

**Table 4.** Diagnostic Efficacy of Biopsy versus Brush Cytology in Biliary Strictures.

| Technique | Patients (n) | SN (95 % CI) | SP (95 % CI) | PPV (95 % CI) | NPV (95 % CI) | Accuracy (95 % CI) |
| --- | --- | --- | --- | --- | --- | --- |
| Forceps Biopsy | 54 | 65.8% (49.4-79.9%) | 92.3% (63.9-99.8%) | 96.4% (81.6-99.9%) | 46.1% (26.5-66.6%) | 72.2% (58.3-83.5%) |
| Brush Cytology | 89 | 56.7% (44.7-68.2%) | 93.3% (68.0-99.8%) | 83.1% (73.7-90.2%) | 30.43% (17.7-45.7%) | 62.9% (52.0-72.9%) |

SN: Sensitivity, SP: Specificity, PPV: Positive Predictive Value, NPV: Negative Predictive Value, Accuracy: Overall diagnostic accuracy.

In summary, while both biopsy and brushing techniques exhibited high specificity, the biopsy method showed marginally superior sensitivity and overall accuracy in the diagnosis of malignant biliary strictures.

## DISCUSSION

Timely and accurate diagnosis of malignant biliary strictures is of paramount importance in clinical practice because of the radical differences in therapeutic approaches between benign and malignant entities. Our study, conducted at one of the largest tertiary care centers in Pakistan, offers a comparative examination of brush cytology and forceps biopsy for the diagnosis of these strictures, specifically within the context of the South Asian population.

Diagnosing biliary strictures using cytological or histological methods can have a significant impact on the management plan for malignancies and benign conditions, and ERCP-based tissue diagnosis can be achieved through brush cytology or intraductal forceps biopsy [8]. Our study demonstrated a sensitivity of 56.7% for brushings and 65.8% for biopsies, suggesting results comparable to those of most previously reported studies. For instance, a study by Pugliese et al. observed a biopsy sensitivity of 53%, almost aligning with a brush sensitivity of 54% [10].The variation in brush sensitivity was also noteworthy, with the lowest being 26% reported by Jailwala et al. and the highest being 72% in Kitajima and colleagues’ analysis, where high-grade dysplasia was considered malignant for sensitivity calculations [11,12]. Weber et al. considered suspicious cells as malignant based on their sensitivity calculations in diagnosing hilar cholangiocarcinoma, yielding a brush sensitivity of 41% and biopsy sensitivity of 53% [13].

Lastly, a meta-analysis completed by Navaneethan et al. reported a pooled sensitivity of 45 (95% CI 40-50%) [8]. A comparison of these numbers highlights the importance of the techniques and criteria adopted for diagnosis. It also underscores the need for continuous advancements in the diagnostic tools and criteria for malignancy assessment. While our study presents findings within the spectrum of prior results, inherent variability remains in brushings and biopsies, emphasizing the importance of contextual interpretation in clinical practice.

However, differences in sensitivity between the two methods should also be explored. Forceps biopsy demonstrated superior sensitivity (65.8%) compared to brush cytology (56.7%). This finding is consistent with previous studies that proposed that forceps biopsy tends to be more sensitive, owing to its capacity to obtain larger and more representative tissue samples, especially in focal strictures [14,15]. This finding may lead to a more definitive diagnosis. In another study comparing different tissue sampling modalities, forceps biopsy had a significantly higher sensitivity than transpapillary brush cytology, and prior balloon dilatation of the stenosis improved the sensitivity, negative predictive value, and accuracy of forceps biopsy [16].

However, brush cytology, although less invasive, may have limitations in diagnosing biliary strictures, especially when the stricture is patchy or when sample collection is inadequate [17]. The use of brush cytology coupled with routine cytology, fluorescence in-situ hybridization (FISH), and next generation sequencing (NGS) has been explored to improve diagnostic sensitivity [18]. FISH coupled with routine cytology appears to be the method of choice for improving the diagnostic sensitivity [19].

The high specificity associated with both diagnostic techniques (93.3% for brushing and 92.3% for biopsy) that we observed was in alignment with prior research, which suggests that a positive result, whether from brushing or biopsy, strongly indicates the presence of malignancy [11–13]. The consistency of this result across both cohorts underscores the reliability of these tests in minimizing false positives. Correctly identifying malignant strictures at an early stage is paramount not only for improving patient outcomes but also for tailoring treatment regimens to prevent unnecessary interventions. Early and accurate diagnosis can lead to timely and targeted treatment, potentially reducing the morbidity and mortality associated with advanced-stage diseases. Furthermore, accurate and early diagnosis is highly cost-effective. It potentially curtails expenses related to prolonged treatments and hospitalizations that may arise from misdiagnoses or delayed interventions. Moreover, reducing the need for repeat diagnostic procedures owing to inconclusive initial results can further alleviate the financial burden on both the healthcare system and patients.

An interesting observation was the change in the post-ERCP diagnosis in both cohorts. There was a decrease in the number of patients diagnosed with cholangiocarcinoma post-ERCP, with an increase in benign biliary stricture diagnoses. This might underscore the importance of integrating clinical, radiological, and histopathological data for definitive diagnosis, as relying solely on preprocedural evaluations can sometimes lead to misleading conclusions.

Our study had certain limitations. Because this was a single-center study, our findings may not be generalizable to a broader population. Moreover, there was a lack of a cohort that underwent both forceps biopsy and brush cytology, which could have been useful for calculating their holistic sensitivity in diagnosing malignant strictures. It would also be beneficial to assess the adverse event rates associated with each technique in future studies, given the known risks associated with forceps biopsy.

## CONCLUSION

In conclusion, both brush cytology and forceps biopsy offer valuable diagnostic insight into biliary strictures. Forceps biopsy demonstrated slightly better diagnostic efficacy in our cohort, with a higher sensitivity and overall accuracy. However, the choice between these two methods should be based on clinical judgment, patient safety, and the available resources. Our findings provide a foundation for further research in Pakistan, aiming for the continual enhancement of diagnostic standards and patient care for biliary strictures.

## Data Availability

All data produced in the present study are available upon reasonable request to the authors

## Acknowledgments

None

